# Tracking the onset date of the community spread of SARS-CoV-2 in Western Countries

**DOI:** 10.1101/2020.04.20.20073007

**Authors:** Edson Delatorre, Daiana Mir, Tiago Gräf, Gonzalo Bello

**Author notes:** Corresponding author: GB. All authors contributed equally to this work. **Sponsorship:** GB is a recipient of CNPq fellowship for Productivity in technological Development and Innovative Extension (Grant 302317/2017-1) and is funded by grant from FAPERJ (Grant E-26/202.896/2018). D.M is member of the Sistema Nacional de Investigadores (National Research System-SNI-ANII, UY).

## Abstract

The SARS-CoV-2 rapidly spread around the world during 2020, but the precise time in which the virus began to spread locally is currently unknown for most countries. Here, we estimate the probable onset date of the community spread of SARS-CoV-2 from the cumulative number of deaths reported during the early stage of the epidemic in Western Europe and the Americas. Our results support that SARS-CoV-2 probably started to spread locally in all western countries analyzed between the middle of January and early February 2020, thus long before community transmission was officially recognized and control measures were implemented.

---

A novel Betacoronavirus, designated SARS-CoV-2, was identified as the causative agent of a severe acute respiratory disease (now known as COVID-19) in Wuhan, Hubei province, China, in December 2019.^(1-2)^ In the following weeks, SARS-CoV-2 rapidly spread around the world, infecting more than 2 million people and causing more than 135,000 deaths as of April 15th, 2020.^(3)^ The exponential growth of the COVID-19 pandemic has overloaded hospitals and governments’ response measures have disrupted social contacts for >1 billion inhabitants worldwide.

The first infections of SARS-CoV-2 identified in Europe and North America in January 2020, were related to travelers returning from China.^(4-5)^ Since February 2020, community transmission of SARS-CoV-2 was already documented in the USA and multiple European countries.^(6)^ However, the precise start time of sustained SARS-CoV-2 local transmission in most of these countries is currently unknown. The presence of a substantial fraction of asymptomatic, but infectious individuals probably facilitated the undocumented dissemination of the novel coronavirus within and among countries before its detection by public health systems.^(7)^ This suggests that when domestic infections with SARS-CoV-2 were reported, epidemics were probably already growing exponentially in most countries for some time.

Genomic epidemiology could be a useful tool to track the geographic spread of SARS-CoV-2 over time and to infer the timing of community transmission. Its implementation has enabled to trace back the time of the most recent common ancestor (T_MRCA_) of SARS-CoV-2 to late November 2019,^(8)^ consistent with epidemiological findings that show viral local transmission in Wuhan by the middle December 2019.^(9)^ Within this same framework, a recent analysis of SARS-CoV-2 genomes from 21 neighborhoods in New York city (NYC) revealed that the NYC epidemic has been mainly sourced from untracked transmission between the USA and Europe and also pointed that the virus was likely circulating in the city weeks before the first confirmed SARS-CoV-2 infection case.^(10)^ At the moment, however, it is challenging to estimate the onset date of local transmission clusters within countries (or cities) based solely on genetic data. Due to the short period of time since the beginning of the outbreak and to the uneven geographic sampling of SARS-CoV-2 genomes, in most cases, there’s still not enough phylogenetic signal to define location-specific clusters.^(8,11)^

Here, we aimed to develop a simple inference method to estimate the onset date of the community spread of SARS-CoV-2 in different countries from the cumulative number of reported deaths during the early stage of the epidemic. The cumulative number of deaths could be a reliable tracker of the SARS-CoV-2 epidemic’s progress within a country.^(12)^ The lack of testing and the great proportion of asymptomatic subjects profoundly restrain the capacity of most countries to count the true number of SARS-CoV-2 infections, while the smaller number of deaths is less prone to such degree of underestimation. Furthermore, the cumulative number of reported deaths represents a time-delayed tracker of the SARS-CoV-2 epidemic and thus provides valuable information on early epidemic dynamics even when data is obtained after implementation of national control measures that reduce the viral spread. Current estimates support an infection fatality ratio of SARS-CoV-2 of around 1% and a median time between infection and death of around three weeks.^(13-14)^

To infer the probable onset date of the community spread of SARS-CoV-2 in a given location, we assumed that: a) as soon as the virus starts spreading locally, the epidemic starts to grow exponentially; b) the cumulative number of deaths starts to increase exponentially 20 days later than the beginning of the epidemic exponential growth; and c) the rate of exponential growth of the number of deaths remains roughly constant during the epidemic early weeks. For this study we selected China and those countries from Western Europe (Belgium, France, Germany, Italy, Netherlands, Spain, United Kingdom [UK]), North America (New York, USA) and South America (Brazil) that were most heavily affected until 5th April 2020.^(3)^ To capture the early period of exponential growth of virus transmission, before national control measures were implemented, we set the start point to the day when the cumulative number of deaths was above one and then counted the cumulative number of deaths during the following three weeks. For Brazil, we used the cumulative number of deaths during the first two weeks because first social distancing measures were taken before detection of the first death. Then, we estimated the number of total infected individuals from the cumulative number of deaths by applying a mean infection fatality rate of 1% and a time-delayed from infection to death of 20 days. Finally, we plotted the projected number of total infected individuals on a logarithmic scale along time and performed a linear regression analysis to estimate the probable onset date of the SARS-CoV-2 community transmission as the X-intercept point of the analysis. Linear regression analyses were performed using Graph Pad v6 (Prism Software, La Jolla, California, USA).

The cumulative number of reported deaths has grown exponentially during the selected period in all countries analyzed with mean estimated epidemic doubling times ranging from 2.6 days to 3.8 days, consistent with previous estimations [Supplementary data (Figure 1)].^(15-17)^ Linear regression analyses of the log number of the projected total infected individuals over time resulted in a high coefficient of determination (r^2^ ≥ 0.95) and significant increase (*P* < 0.0001) for all analyzed countries (Figure 1). To validate our approach, we first estimated the beginning of community transmission of SARS-CoV-2 in locations with previous molecular clock estimates. Using our framework, we traced the onset date of community transmission of SARS-CoV-2 to early December 2019 in China and to early February 2020 in New York (Figure 1 and Table 1). These estimates were consistent with molecular clock calibrations from SARS-CoV-2 genomic data (Table 1)^(8,10)^, and suggest that both approaches might provide roughly similar results.

**Table 1.**
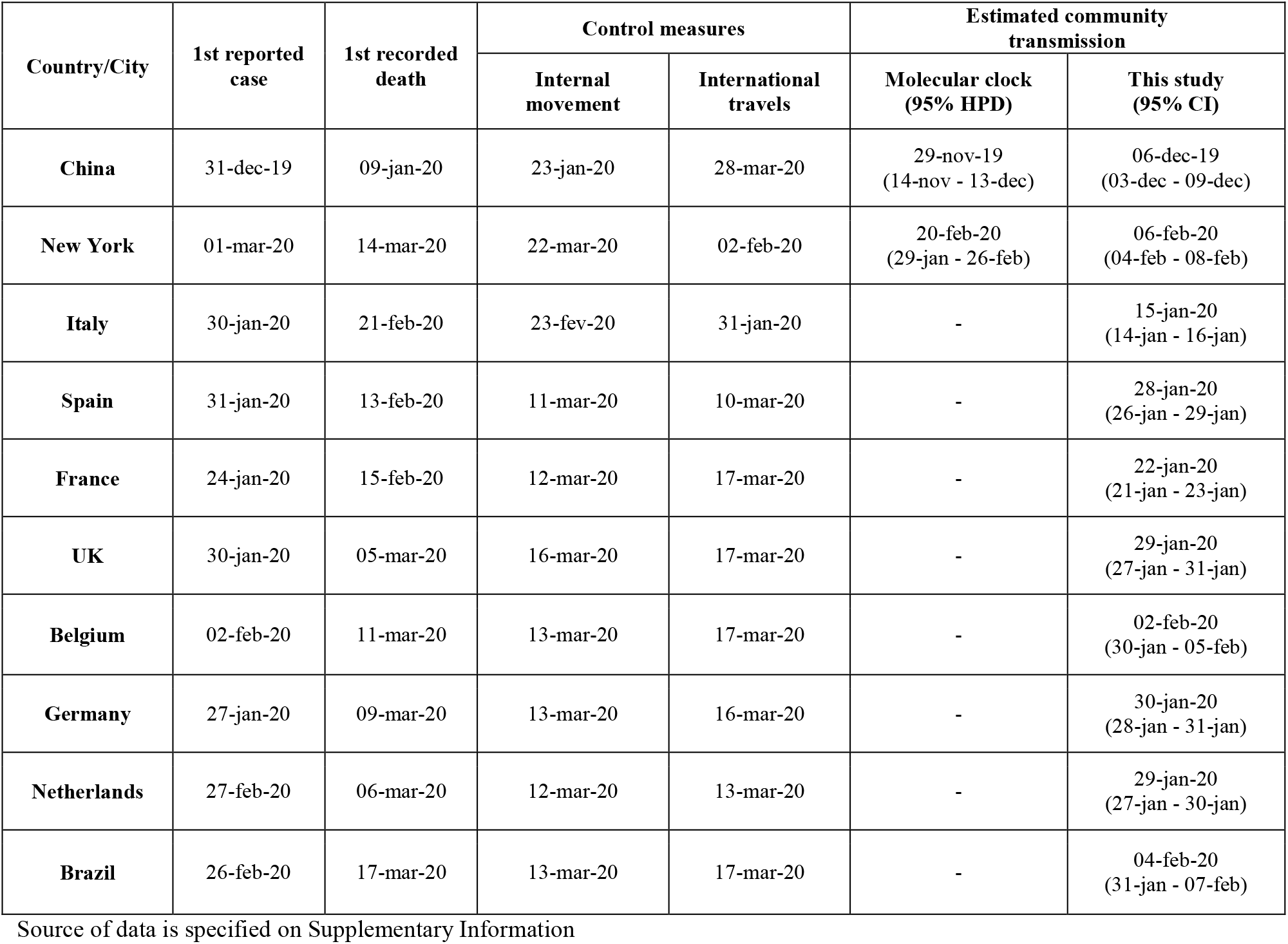
COVID-19 epidemiologic timeline and implementation of related control measures in selected countries.

| Country/City | 1st reported case | 1st recorded death | Control measures |  | Estimated community transmission |  |
| --- | --- | --- | --- | --- | --- | --- |
|  |  |  | Internal movement | International travels | Molecular clock (95% HPD) | This study (95% CI) |
| **China** | 31-dec-19 | 09-jan-20 | 23-jan-20 | 28-mar-20 | 29-nov-19<br>(14-nov - 13-dec) | 06-dec-19<br>(03-dec - 09-dec) |
| **New York** | 01-mar-20 | 14-mar-20 | 22-mar-20 | 02-feb-20 | 20-feb-20<br>(29-jan - 26-feb) | 06-feb-20<br>(04-feb - 08-feb) |
| **Italy** | 30-jan-20 | 21-feb-20 | 23-fev-20 | 31-jan-20 | - | 15-jan-20<br>(14-jan - 16-jan) |
| **Spain** | 31-jan-20 | 13-feb-20 | 11-mar-20 | 10-mar-20 | - | 28-jan-20<br>(26-jan - 29-jan) |
| **France** | 24-jan-20 | 15-feb-20 | 12-mar-20 | 17-mar-20 | - | 22-jan-20<br>(21-jan - 23-jan) |
| **UK** | 30-jan-20 | 05-mar-20 | 16-mar-20 | 17-mar-20 | - | 29-jan-20<br>(27-jan - 31-jan) |
| **Belgium** | 02-feb-20 | 11-mar-20 | 13-mar-20 | 17-mar-20 | - | 02-feb-20<br>(30-jan - 05-feb) |
| **Germany** | 27-jan-20 | 09-mar-20 | 13-mar-20 | 16-mar-20 | - | 30-jan-20<br>(28-jan - 31-jan) |
| **Netherlands** | 27-feb-20 | 06-mar-20 | 12-mar-20 | 13-mar-20 | - | 29-jan-20<br>(27-jan - 30-jan) |
| **Brazil** | 26-feb-20 | 17-mar-20 | 13-mar-20 | 17-mar-20 | - | 04-feb-20<br>(31-jan - 07-feb) |

Source of data is specified on Supplementary Information

**Figure 1.**
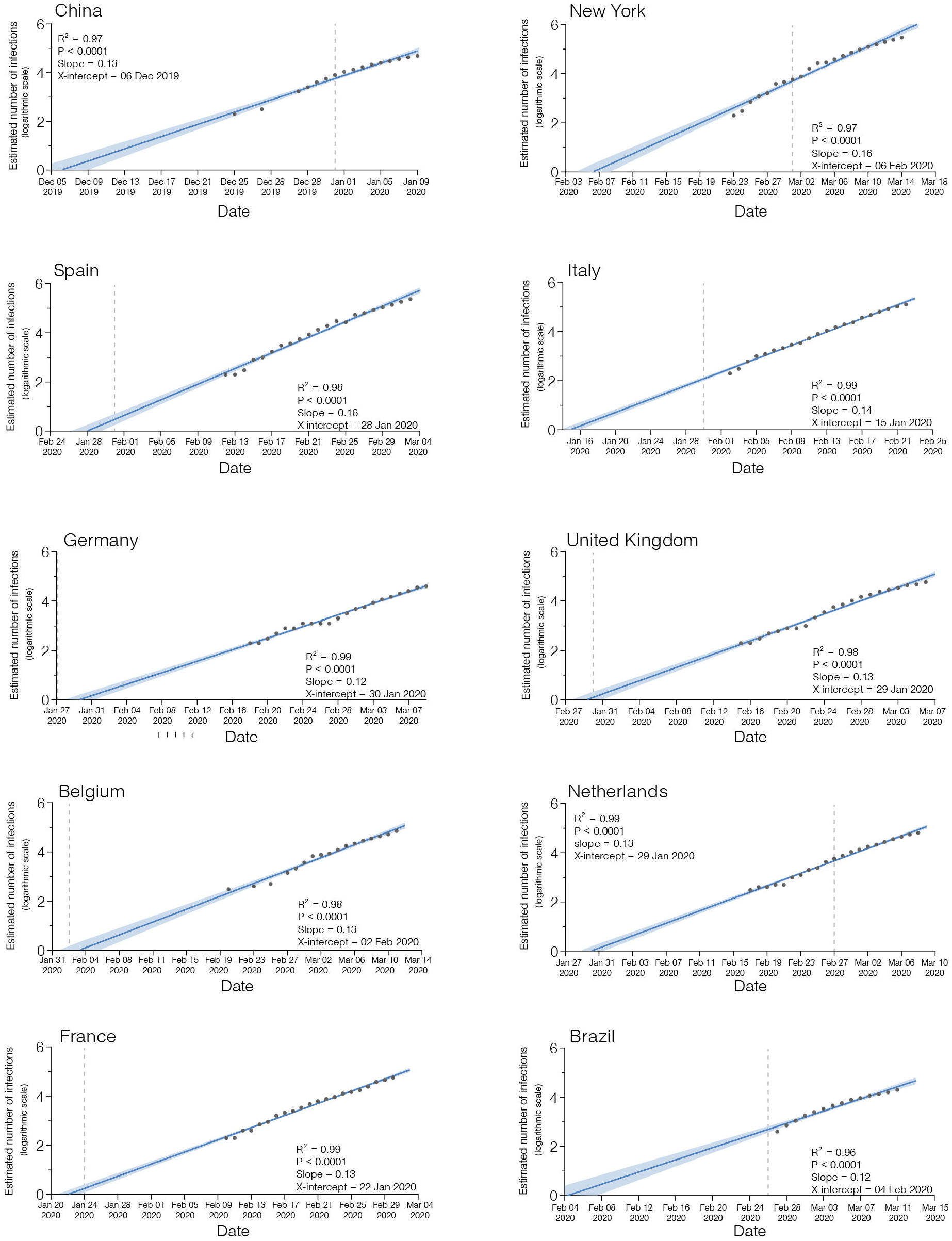
Linear regressions of the log-transformed estimated number of total infections by SARS-CoV-2 over time in selected countries. The light blue shadow represents the 95% confidence intervals of the log-linear regression line (dark blue) fitted on the estimated number of infections (gray dots) for each country as indicated. The goodness of fit (R^2^), *P*-value, slope and X-intercept for each country are presented for each graph. The date of the 1^st^ reported SARS-CoV-2 case in each country is indicated by the gray dashed line.

We next used our approach to estimate the onset date of local transmission of SARS-CoV-2 in Western Europe. Our findings suggest that community transmission might have started around middle January 2020 in Italy and between late January and early February 2020 in the remaining countries (Figure 1). In some countries (Italy and Netherlands) community transmission was traced long before (2-4 weeks) the first confirmed SARS-CoV-2 infection case; while in others (Spain, France, United Kingdom, Germany, and Belgium) the onset date roughly coincides with the time of detection of the first imported cases (Figure 1). In all countries analyzed, however, SARS-CoV-2 probably started to spread locally long before community transmission was officially recognized and control measures for social distancing and air travel restrictions were implemented (Table 1). Of note, our estimation of the onset date of community transmission of SARS-CoV-2 in European countries is consistent with the estimated T_MRCA_ of some European and European/American clades like the A1a (28^th^ January: 19^th^ January - 17^th^ February) and the A2a (25^th^ January: 19^th^ January - 2^nd^ February).^(10)^ These results suggest that early community transmission of SARS-CoV-2 in Europe could have been fueled by the rapid dissemination of few regional clades.

South America was one of the last geographic regions in the world to declare community transmission of SARS-CoV-2. Our study, however, supports that the virus started to spread in Brazil at around the early February 2020 (Figure 1), thus coinciding with the epidemic expansion in North America and Western Europe and preceding by more than 20 days the first confirmed SARS-CoV-2 infection case in the country (Table 1). According to a recent report of the InfoGripe (http://info.gripe.fiocruz.br) - a system of the Fundação Oswaldo Cruz (Fiocruz) that weekly monitor the number of hospitalizations of patients who have had symptoms such as fever, cough or sore throat and difficulty breathing in Brazil - the number of such cases started to increase above levels observed in 2019 since the middle February 2020.^(18)^ More importantly, of the retrospective cases that tested positive for some respiratory virus, nine cases were identified as SARS-CoV-2 in the eight epidemiological week, between 16th and 22nd February (https://covid.saude.gov.br/). These epidemiological data fully agree with our estimation and clearly support that SARS-CoV-2 was already circulating in the Brazilian population for some weeks before being detected on 26th February 2020.

Our findings support that, except Italy, SARS-CoV-2 started to spread locally in several countries from Western Europe and the Americas at around the same time. Despite this, early epidemiological dynamics seem to be quite different across countries. Among those countries where SARS-CoV-2 started to spread in late January, Spain passed the milestone of 1,000 deaths earlier (20th March) than France (24th March), the UK (27th March), Netherlands (31th March), and Germany (3rd April).^(3)^ Similarly, among those regions where SARS-CoV-2 started to spread in early February, New York (30th March) and Belgium (2nd April) reached 1,000 deaths much earlier than Brazil (10th April).^(3)^ Some studies support that mortality rate might vary across regions according to the availability of health-care facilities, underlying demography and environmental factors (average temperature, precipitation seasonality and levels of air pollution).^(13,19-22)^ Early dynamics in the reported number of deaths may also be influenced by the time when local social distancing measures were implemented (Table 1) and by the percentage of deaths with laboratory confirmation within each country.

The narrow confidence intervals of our estimates should be interpreted with caution because our method does not account for uncertainty and geographic variability in the mortality rate and lag-time between infection and death.^(13-14)^ As more clinical and epidemiological data becomes available, it will be possible to refine these estimates by using more accurate regionally-specific parameters. Despite this, data from countries that implement wide-scale testing for SARS-CoV-2 since the beginning of the outbreak, including people who have mild or no symptoms, supports that the mortality rate will probably not exceed 1% of the total number of infected individuals.^(23)^ Hence, the estimates presented here should be closer to the upper limit of the likely time range when virus community transmission has started. Another limitation is that our method is sensitive to underestimation of the true number of deaths from SARS-CoV-2 and should thus be used with caution in countries with limited capacity for testing and/or substantially delayed in the report of deaths.

In summary, our results support that community transmission of SARS-CoV-2 probably started in many western countries between middle January to early February 2020, thus long before control measures to restrict air travels and promote social distancing were implemented. That quite long period of cryptic community transmission (> 4 weeks) in all analyzed countries draws attention to the great challenge of tracking the early global spread of SARS-CoV-2 and supports that control measures should be adopted at least as soon as first imported cases are detected in a new geographic region. This is especially important in the light of studies showing that SARS-CoV-2 very likely will enter in a regular circulation after the initial pandemic wave, causing recurrent outbreaks in the next years whose frequency and intensity are dependent upon virus’ biological features still not well understood, like the duration of immunity that SARS-CoV-2 can induce.^(24)^ In this scenario, intense virological surveillance is pivotal to early detect the virus re-emergence, informing systems of contact tracing and providing evidence to carry out appropriate control measures.

## Data Availability

Data sharing is not applicable to this article as no new data were created or analyzed in this study.

